# Factors Associated with Predischarge Morbidity and Mortality in Small for Gestational Age Infants: A Single-Center Retrospective Cohort Study

**DOI:** 10.64898/2026.10.07.26364995

**Authors:** Christopher Cinkus-Rios, Samantha Pickell, Charlotte Thompson, Miles Bomback, Alex Lyford, Thomas Hays

## Abstract

**Objective:** To identify factors associated with predischarge morbidity and mortality among small for gestational age infants.

**Study Design:** We conducted a retrospective single-center cohort study of small for gestational age infants admitted to a level IV neonatal intensive care unit. We determined the association of clinical characteristics, congenital anomalies, and genetic disorders with the primary outcome of predischarge morbidity or mortality and characterized morbidity and mortality across congenital anomaly subgroups.

**Results:** Among 293 small for gestational age infants, 18 (6.1%) had a genetic disorder and 103 (35.2%) had a major congenital anomaly, with congenital heart disease (CHD) representing the largest subgroup. The primary outcome of morbidity or mortality occurred in 93 infants (31.7%); 74 infants (25.3%) experienced morbidity and 19 (6.5%) died. CHD (adjusted odds ratio [aOR]: 16.5, 95% CI 6.44–45.7) and multiple congenital anomalies (aOR: 17.8, 95% CI 4.64–71.1) were found to be strongly associated with morbidity or mortality, after accounting for sex, maternal race, and intubation > 7 days. Statistically significant associations were also found with the presence of musculoskeletal anomalies as well as genetic disorders in infants without congenital anomalies, though in limited sample sizes. CHD was the largest contributor to morbidity, whereas multiple congenital anomalies accounted for the greatest proportion of mortality. Infants with abdominal wall defects and central nervous system anomalies experienced no morbidity or mortality.

**Conclusion:** Congenital heart disease and multiple congenital anomalies were strongly associated with morbidity and mortality among SGA infants, whereas the degree of SGA alone was not. These findings suggest that risk is not explained by the degree of SGA alone and may be more strongly associated with congenital disease and other underlying factors.

**Key points:**

- Risk is highly variable in SGA infants
- CHD predicts adverse outcomes
- Multiple anomalies predict adverse outcomes
- Degree of SGA alone does not explain risk

## Introduction

Small for gestational age (SGA) is defined as birth weight <10^th^ percentile for gestational age based on population reference curves.^1^ SGA often follows fetal growth restriction (FGR), which refers to failure of a fetus to reach its healthy potential growth, typically defined as <10^th^ percentile for similar fetal growth curves.^2^ Many factors can cause FGR and subsequent SGA birth. Placental insufficiency is the most frequently identified cause.^3^ Rarely, FGR results from intrauterine infections, such as cytomegalovirus, rubella, toxoplasmosis, varicella, and syphilis.^2^

The diagnosis of SGA does not distinguish between an infant that is constitutionally small but healthy versus one that failed to reach their innate potential birth weight. Consistent with this, the outcomes of SGA infants are highly variable. SGA infants experience mortality rates 2–3 times higher than appropriate for gestational age (AGA) infants, however, the cause(s) of this are incompletely understood.^4,5^ In the setting of FGR, fetuses often accumulate lower stores of glycogen and adipose.^6^ Chronic placental insufficiency can disrupt fetal hematopoiesis by shifting production away from platelets and white blood cells to erythropoiesis.^7^ SGA infants also have greater heat loss due to an increased surface area-to-volume ratio. Because of these characteristics, SGA infants broadly have increased risk of neonatal metabolic and hematologic complications, including hypoglycemia, hypothermia, polycythemia, thrombocytopenia, leukopenia, as well as sepsis and death.^4,8–12^ Yet it remains difficult to predict which will experience neonatal morbidity or mortality.^4,12^

Increasingly, genetic disorders are identified as underlying SGA and FGR.^12,13^ Genetic disorders are approximately twice as common in SGA infants compared to AGA infants.^12,15^ Additionally, SGA infants frequently bear copy number variants (CNVs), with 10-16% of SGA infants having CNVs that are pathogenic or likely pathogenic.^14–16^ A large number of single gene disorders have also been found to cause SGA.^13^ In the setting of congenital anomalies, infants are more likely to be SGA.^13^ This association is not fully understood, although underlying genetic disorders may contribute to both low birth weight and congenital anomalies. SGA infants with genetic disorders are more than twice as likely to experience morbidity and mortality.^12^

Despite the well-recognized risks associated with SGA, neonatal outcomes remain highly heterogeneous. Some SGA infants experience an uncomplicated clinical course, whereas others develop severe morbidity or die despite having a similar degree of SGA. Birth weight alone does not adequately explain this variability, suggesting that underlying factors may contribute importantly to neonatal risk. Better understanding of these factors could improve early risk stratification and clinical management. Therefore, we sought to characterize factors associated with predischarge morbidity and mortality in SGA infants, with particular attention to congenital anomalies and genetic disorders.

## Materials and Methods

### Study subjects and data collection

This study was approved by the Columbia University institutional review board (protocol AAAT2607). We performed a retrospective single-center cohort study at a level IV neonatal intensive care unit (NICU). We included SGA infants (defined by Fenton criteria),^17^ admitted from June 2023 through April 2025, born after at least 22 weeks’ gestation. We excluded infants transferred from other institutions to limit ascertainment bias.^18^ We excluded cases with chromosomal trisomies.

We conducted a manual review of electronic health records and abstracted the following data: antenatal corticosteroid treatment, birth weight, gestational age, sex, delivery mode, presence of major congenital anomalies or diagnosed genetic disorders, vasopressor or inotrope treatment, endotracheal intubation duration, maternal age, race, and ethnicity, and the occurrence of morbidity or mortality prior to NICU discharge. Morbidities consisted of the following: acute kidney injury (AKI; defined as clinician denoted “oliguria,” “anuria,” or “acute kidney injury”); intraventricular hemorrhage (IVH; defined as grade II or greater by Papile classification^19^); necrotizing enterocolitis (NEC; defined as stage II or greater by Bell staging^20^); bronchopulmonary dysplasia (BPD; defined as grade 2 or greater by Jensen criteria^21^); and shock defined as need for treatment with a vasopressor or inotrope. Major congenital anomalies were grouped by organ system. Birth weight percentiles and z-scores were determined using Fenton criteria.^17^

### Statistical Analyses

We performed a Firth penalized logistic regression to determine factors associated with morbidity or mortality. The sample size consisted of all eligible infants during the study period; no a priori sample size was performed. The primary outcome was the composite of either morbidity or mortality prior to NICU discharge. Exposures of interest consisted of congenital anomaly subtype, with no anomaly as reference, the presence of known genetic disorders, mode of delivery (vaginal delivery as reference), gestational age, severe SGA (defined as birth weight < 3^rd^ percentile for gestational age), antenatal corticosteroid exposure (any treatment versus no exposure), and the presence of abnormal umbilical artery Doppler velocimetry findings (documented abnormal findings versus normal findings or no documentation of Doppler studies).^5^ Associations for these exposures were estimated separately for infants with and without congenital anomalies. Models additionally adjusted for sex (female as reference), maternal self-identified race (non-White versus White), and intubation > 7 days; as above, associations for these covariates were estimated separately for infants with and without congenital anomalies. Cases with central nervous system anomalies and abdominal wall defects (n=11) were not included as individual congenital anomaly categories in the anomaly subtype analysis because no infants in these groups experienced morbidity or mortality; these infants remained included in all other analyses.

We also characterized patterns of morbidity and mortality with alluvial diagrams.^22^ Alluvial diagrams visualize events within groups of individuals represented by bands proportional in size to the number of individuals to highlight patterns.^22^ We grouped infants by organ system of major congenital anomalies. Infants were then categorized based on occurrence of morbidity and mortality. Analyses and data visualization were performed using R (Version 4.5.2). Details regarding the scripts and packages used in this project can be found in **Supplemental Methods.**

## Results

### Cohort description

The cohort consisted of 293 SGA infants. Most had severe SGA, with 222 (75.8%) below the 3rd percentile and 156 (53.2%) below the 1st percentile for birth weight. The primary outcome of predischarge morbidity or mortality was reached in 93 (31.7%) infants, of which 74 (25.3%) experienced morbidity and 19 (6.5%) died (**Table 1**). Abnormal fetal Dopplers were identified in 146 of 293 (49.8%); in 18 cases (6.1%), no assessment of fetal Dopplers was documented.

**Table 1.**
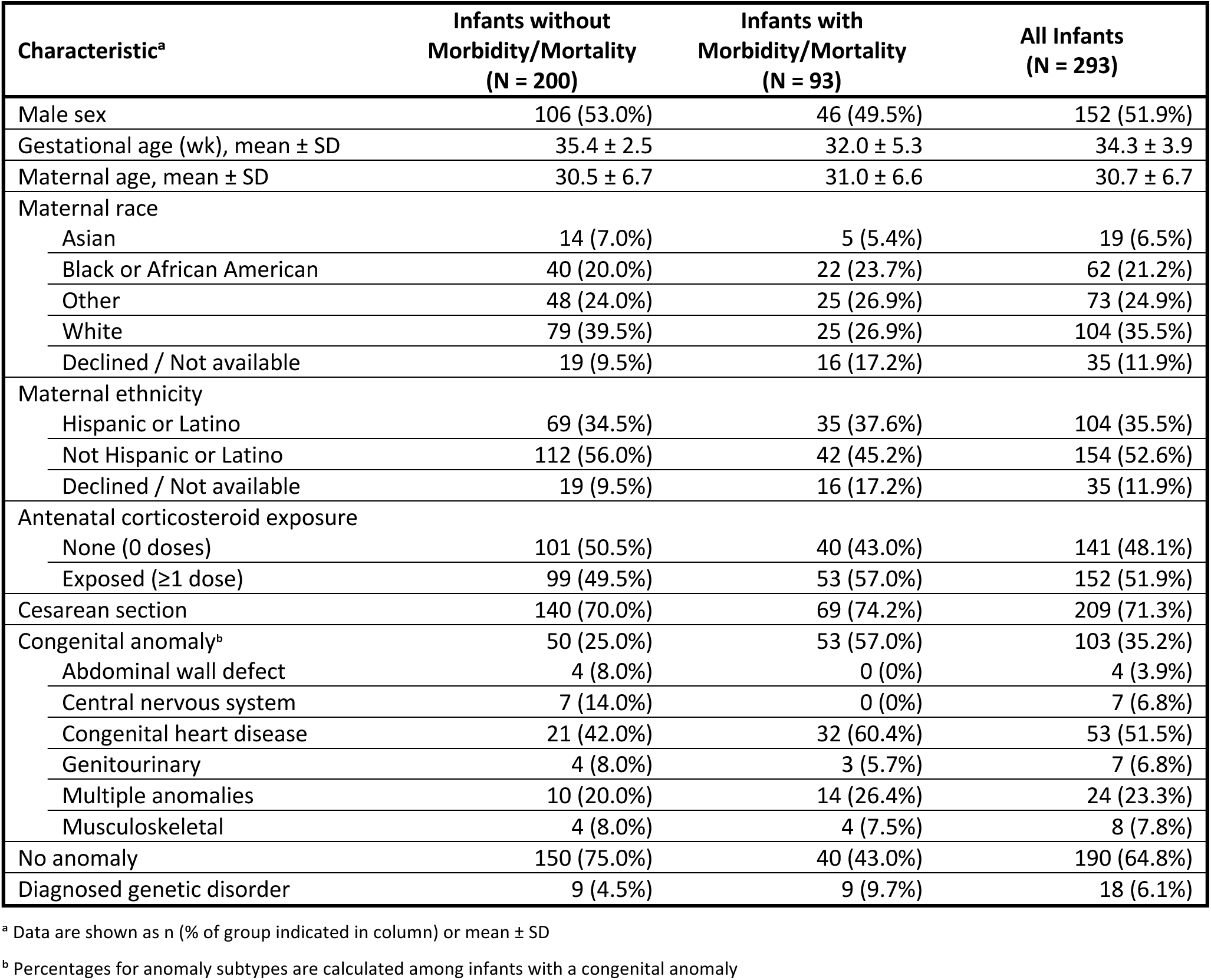
Characteristics of Cohort.

| Characteristic<sup>a</sup> | Infants without<br>Morbidity/Mortality<br>(N = 200) | Infants with<br>Morbidity/Mortality<br>(N = 93) | All Infants<br>(N = 293) |
| --- | --- | --- | --- |
| Male sex | 106 (53.0%) | 46 (49.5%) | 152 (51.9%) |
| Gestational age (wk), mean ± SD | 35.4 ± 2.5 | 32.0 ± 5.3 | 34.3 ± 3.9 |
| Maternal age, mean ± SD | 30.5 ± 6.7 | 31.0 ± 6.6 | 30.7 ± 6.7 |
| Maternal race |  |  |  |
| Asian | 14 (7.0%) | 5 (5.4%) | 19 (6.5%) |
| Black or African American | 40 (20.0%) | 22 (23.7%) | 62 (21.2%) |
| Other | 48 (24.0%) | 25 (26.9%) | 73 (24.9%) |
| White | 79 (39.5%) | 25 (26.9%) | 104 (35.5%) |
| Declined / Not available | 19 (9.5%) | 16 (17.2%) | 35 (11.9%) |
| Maternal ethnicity |  |  |  |
| Hispanic or Latino | 69 (34.5%) | 35 (37.6%) | 104 (35.5%) |
| Not Hispanic or Latino | 112 (56.0%) | 42 (45.2%) | 154 (52.6%) |
| Declined / Not available | 19 (9.5%) | 16 (17.2%) | 35 (11.9%) |
| Antenatal corticosteroid exposure |  |  |  |
| None (0 doses) | 101 (50.5%) | 40 (43.0%) | 141 (48.1%) |
| Exposed ($\geq 1$ dose) | 99 (49.5%) | 53 (57.0%) | 152 (51.9%) |
| Cesarean section | 140 (70.0%) | 69 (74.2%) | 209 (71.3%) |
| Congenital anomaly<sup>b</sup> | 50 (25.0%) | 53 (57.0%) | 103 (35.2%) |
| Abdominal wall defect | 4 (8.0%) | 0 (0%) | 4 (3.9%) |
| Central nervous system | 7 (14.0%) | 0 (0%) | 7 (6.8%) |
| Congenital heart disease | 21 (42.0%) | 32 (60.4%) | 53 (51.5%) |
| Genitourinary | 4 (8.0%) | 3 (5.7%) | 7 (6.8%) |
| Multiple anomalies | 10 (20.0%) | 14 (26.4%) | 24 (23.3%) |
| Musculoskeletal | 4 (8.0%) | 4 (7.5%) | 8 (7.8%) |
| No anomaly | 150 (75.0%) | 40 (43.0%) | 190 (64.8%) |
| Diagnosed genetic disorder | 9 (4.5%) | 9 (9.7%) | 18 (6.1%) |

<sup>a</sup> Data are shown as n (% of group indicated in column) or mean ± SD
<sup>b</sup> Percentages for anomaly subtypes are calculated among infants with a congenital anomaly

Major congenital anomalies were present in 103 infants (35.2%). Congenital heart disease (CHD) was the most common anomaly subtype (53 infants, 51.5% of anomalous infants), followed by multiple congenital anomalies (24, 23.3%), musculoskeletal anomalies (8, 7.8%), central nervous system anomalies (7, 6.8%), genitourinary anomalies (7, 6.8%), and abdominal wall defects (4, 3.9%). Genetic disorders were diagnosed in 18 infants (6.1%). Birth weight distributions were similar among infants with and without diagnosed genetic disorders (**Figure 1**).

**Figure 1.**
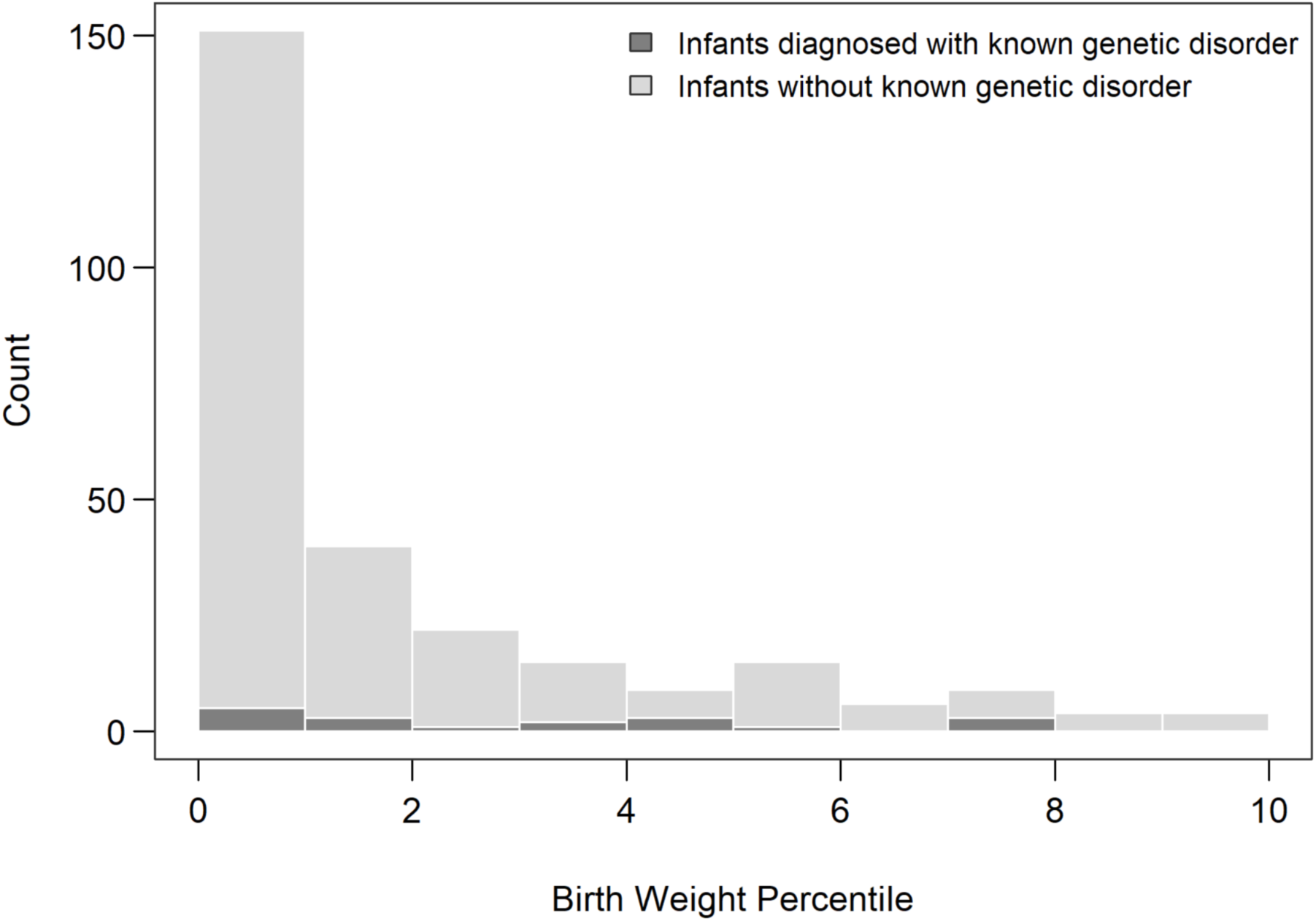
Distribution of birth weight in cohort. Birth weights of individuals (N = 293) were classified as percentiles for gestational age and sex, and infants were stratified based on the presence of diagnosed genetic disorders. Bars represent counts of infants within each percentile-point interval of birth weight. Dark gray indicates infants with known genetic disorders.

### Clinical characteristics distinguishing high-risk SGA infants

In multivariable analysis, CHD, multiple congenital anomalies, and musculoskeletal anomalies were associated with increased odds of morbidity or mortality. Among infants without congenital anomalies, a diagnosed genetic disorder was also associated with increased odds of the composite outcome. Greater gestational age was associated with lower odds of morbidity or mortality in both infants with and without congenital anomalies. Antenatal corticosteroid exposure was associated with lower odds of morbidity or mortality among infants with anomalies (**Figure 2**). Models were adjusted for sex, maternal race, and intubation > 7 days.

**Figure 2.**
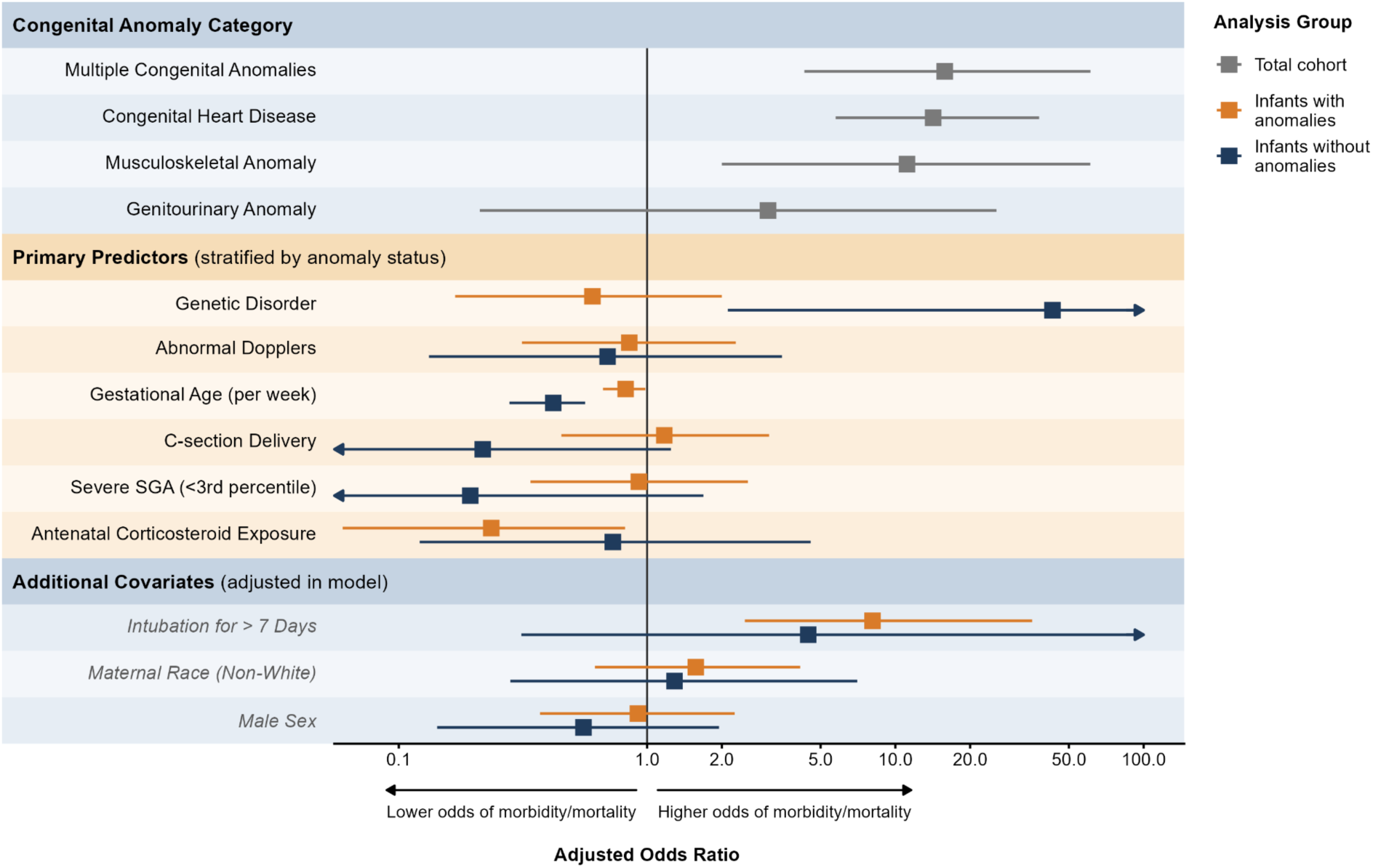
Exposures associated with morbidity or mortality. Forest plot of adjusted odds ratios (aORs) and 95% confidence intervals from Firth penalized logistic regression (n = 293). Congenital anomaly categories (grey) were estimated in the total cohort relative to infants without anomalies. Primary predictors are shown separately for infants with (orange) and without (navy) congenital anomalies, estimated from a model including interactions with anomaly status; male sex, non-White maternal race, and intubation >7 days were included as adjustment covariates and are shown for completeness. Arrows indicate confidence intervals extending beyond the displayed range.

### Patterns of morbidity differed by congenital anomaly subtype

The adverse outcomes in infants grouped by type of congenital anomaly are shown in **Figure 3**. Infants with CHD experienced a high burden of morbidity, frequently consisting of shock. Infants with multiple congenital anomalies disproportionately died, often following acute kidney injury. Infants with genitourinary anomalies often experienced acute kidney injury, but rarely mortality. Infants with musculoskeletal anomalies experienced IVH, BPD, or mortality. In contrast, infants with central nervous system anomalies and abdominal wall defects survived to discharge without major morbidity.

**Figure 3.**
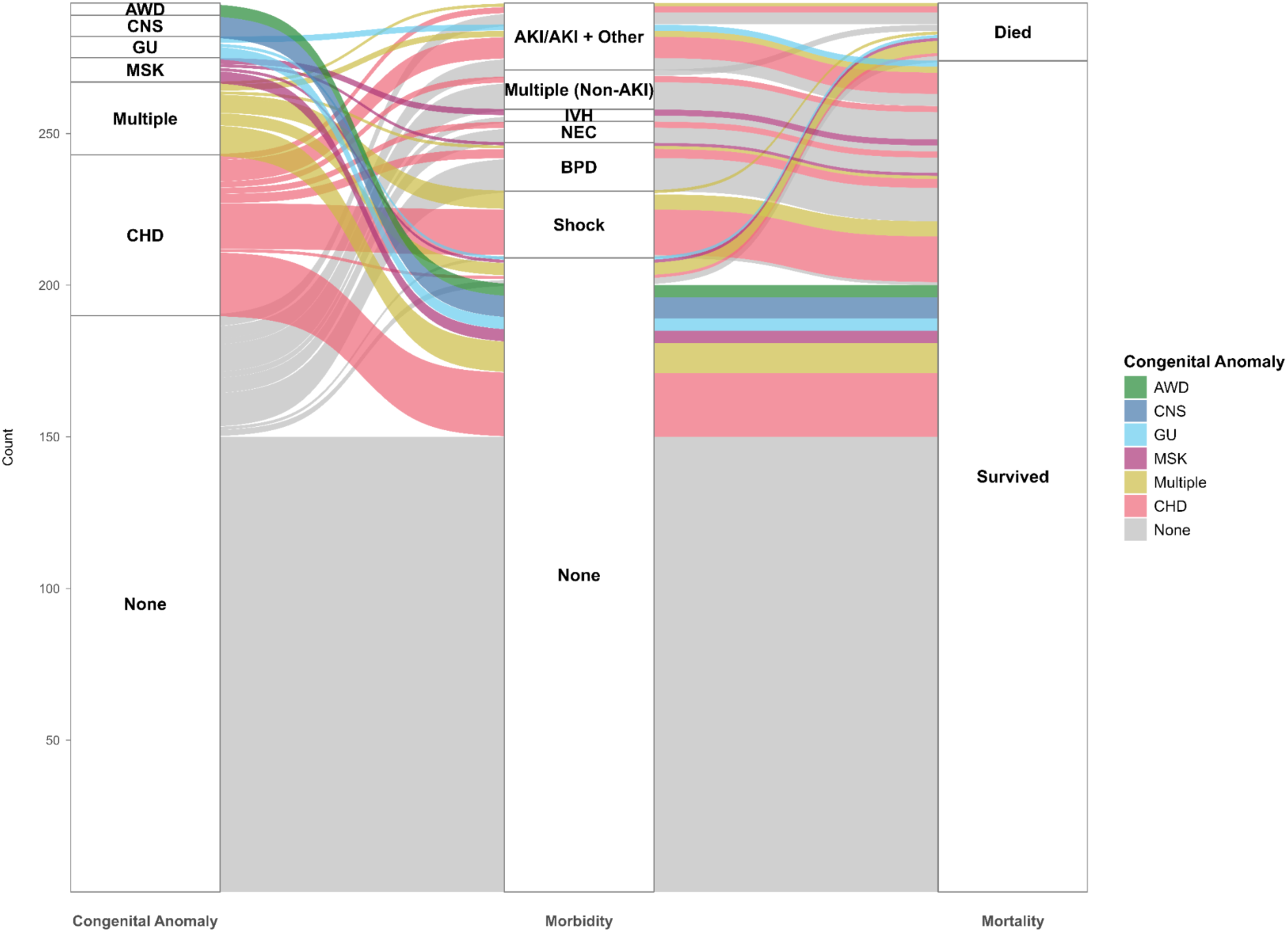
Alluvial diagram depicting the relationship between congenital anomalies, morbidity, and mortality. Infants were grouped based on presence of major congenital anomalies (organ systems shown at left), the occurrence of morbidities (middle), and survival to NICU discharge (right). Band width is proportional to the number of infants. Abbreviations: AKI (acute kidney injury), AWD (abdominal wall defect), BPD (bronchopulmonary dysplasia), CHD (congenital heart disease), CNS (central nervous system), GU (genitourinary), IVH (intraventricular hemorrhage), MSK (musculoskeletal), and NEC (ncrotizing enterocolitis).

Notably, adverse outcomes were not confined to infants with congenital anomalies. A subset of infants without major congenital anomalies experienced acute kidney injury, intraventricular hemorrhage, necrotizing enterocolitis, bronchopulmonary dysplasia, or multiple morbidities.

## Discussion

The principal finding of this study is that risk in NICU-admitted SGA infants is highly heterogeneous and is not explained by the severity of SGA alone. CHD, multiple congenital anomalies, and musculoskeletal anomalies were most strongly associated with morbidity or mortality. Genetic disorders were also strongly associated with morbidity or mortality, but only in infants without congenital anomalies. In contrast, severity of SGA was not independently associated with adverse outcomes. Importantly, morbidity and mortality were not confined to infants with congenital anomalies. A subset of non-anomalous SGA infants experienced major morbidity or mortality, suggesting that clinically important heterogeneity persists beyond the presence of congenital anomalies. Together, these findings support viewing SGA as a heterogeneous clinical phenotype in which neonatal risk is not adequately captured by birth weight percentile or the presence of a recognized congenital or genetic disorder alone.

Several specific patterns emerged from our regression and alluvial analyses. Mortality in this cohort was frequently preceded by AKI, while infants with CHD or multiple congenital anomalies frequently experienced shock. In contrast, NEC and BPD occurred predominantly in infants without congenital anomalies. CHD, multiple congenital anomalies, musculoskeletal anomalies, and genetic disorders in infants without congenital anomalies were each independently associated with increased odds of the composite outcome of morbidity or mortality after controlling for sex, maternal race, and prolonged intubation.

The associations found in infants with multiple anomalies and CHD were relatively robust, as they were driven by relatively large sample sizes. Fourteen of 24 infants with multiple anomalies and 32 of 53 infants with CHD experienced morbidity or mortality respectively. In contrast, one of two non-anomalous infants with genetic disorders and four of eight infants with musculoskeletal anomalies experienced morbidity or mortality respectively. We therefore refrain from drawing further conclusions from these latter associations. Notably, genetic testing was not uniform in this cohort and infants may have harbored undiagnosed disorders. Finally, infants with CNS anomalies and abdominal wall defects experienced no observed morbidity or mortality.

Interestingly, the presence of abnormal umbilical artery Doppler findings was not associated with postnatal morbidity or mortality in either SGA infants with or without congenital anomalies. Abnormal umbilical artery Doppler findings have a well-established association with adverse perinatal outcomes.^5^ However, prior studies indicate that the association with postnatal morbidity and mortality may be partially explained by prematurity and antenatal corticosteroid exposure.^23,24^ Furthermore, fetuses with severely abnormal Doppler findings may have experienced stillbirth or termination. Thus, in our analysis, which accounted for gestational age and other factors, the presence of abnormal Dopplers may not have been independently associated with adverse outcomes.

Notably, a subset of non-anomalous infants also experienced morbidity and mortality. Although most infants who experienced morbidity survived, mortality was often preceded by AKI or multiple morbidities. This heterogeneity among non-anomalous SGA infants may reflect differences in the underlying pathogenesis of SGA. Some infants may have been constitutionally small, whereas others may have experienced pathologic FGR related to placental dysfunction or other disruptions in fetal development, placing them at greater risk for adverse neonatal outcomes.

Interestingly, the degree of SGA did not independently distinguish infants who experienced adverse outcomes. This was notable given that more than three-quarters of the cohort had birth weights below the 3^rd^ percentile and more than half were below the 1^st^ percentile. Thus, even within a predominantly severely SGA cohort, smaller birth weight alone did not identify the infants at greatest risk. These findings further suggest that neonatal risk among SGA infants reflects underlying pathophysiology rather than the degree of SGA alone.

Our findings have several implications for clinical practice. They support risk stratification of infants beyond the degree of SGA.^25,26^ The presence of CHD and multiple congenital anomalies may identify infants at elevated risk for adverse outcomes. The significance of our findings regarding musculoskeletal anomalies and genetic disorders is less clear given limited sample sizes. However, morbidity and mortality among infants without congenital anomalies highlight the limitations of identifiable clinical risk factors. Distinguishing constitutionally small infants from those with pathologic FGR remains an important clinical challenge.^27^ Improved identification of pathologic FGR may help identify infants at risk for adverse outcomes when overt risk factors are absent. Prospective multicenter studies incorporating genomic testing, placental pathology, and longitudinal neonatal outcomes may help improve prognostication in SGA infants, particularly those without congenital anomalies.

Our study has notable strengths and weaknesses. It consisted of a detailed manual chart review, enabling identification of granular details regarding congenital anomalies and other characteristics of infants. Complementary regression and alluvial analyses demonstrated risk in SGA infants with congenital anomalies as well as heterogeneity of outcomes in non-anomalous SGA infants. The retrospective single-center design carries risk of selection bias and residual confounding. Findings from a level IV NICU may not generalize to lower acuity settings. The small cohort size limited statistical power within certain anomaly subgroups, particularly CNS and abdominal wall anomalies. Survival bias is possible, as severely growth-restricted fetuses may not have survived to birth. Additionally, placental pathology and antenatal measures of FGR were not uniformly available, limiting our ability to distinguish constitutionally small infants from those with pathologic fetal growth restriction.

### Conclusions

In conclusion, among NICU-admitted SGA infants, morbidity and mortality were strongly associated with CHD and multiple anomalies and possibly associated with musculoskeletal anomalies and with genetic disorders among non-anomalous infants. Morbidity and mortality were not found to be associated with the severity of SGA. Adverse outcomes also occurred in a subset of infants without congenital anomalies, highlighting the clinical heterogeneity within the SGA population. Future studies integrating placental, genomic, and physiologic data may help identify the biologic factors underlying this heterogeneity and refine neonatal risk stratification.

## Ethics Declarations

### Conflicts of interest

none declared

### Ethics approval

The Columbia University Institutional Review Board approved this study (protocol AAAT2607). The need for informed consent was waived given the nature of the de-identified, retrospective cohort. The study was performed in accordance with the Declaration of Helsinki.

### Data Availability

The data underlying this study are not publicly available because they contain information that could compromise participant privacy. De-identified data are available from the corresponding author upon reasonable request and subject to institutional approval.

### Funding

This work was supported by a Precision Medicine Pilot Grant from the Irving Institute via the National Center for Advancing Translational Sciences (UL1TR001873 to TH), and by the Eunice Kennedy Shriver National Institute of Child Health and Human Development (K23HD113827 to TH).

## Notes

### Competing Interest Statement

The authors have declared no competing interest.

